# Hepcidin regulation in Kenyan children with severe malaria and non-typhoidal *Salmonella* bacteremia

**DOI:** 10.1101/2021.04.08.21255042

**Authors:** Kelvin M. Abuga, John Muthii Muriuki, Sophie M. Uyoga, Kennedy Mwai, Johnstone Makale, Reagan Mogire, Alex W. Macharia, Shebe Mohammed, Salim Mwarumba, Neema Mturi, Philip Bejon, J. Anthony G. Scott, Manfred Nairz, Thomas N. Williams, Sarah H. Atkinson

## Abstract

**Background:** Severe malaria and invasive non-typhoidal *Salmonella* (NTS) are life-threatening infections that often co-exist in African children. The iron-regulatory hormone hepcidin is highly upregulated during malaria and controls the availability of iron, a critical nutrient for bacterial growth, within the *Salmonella*-containing vacuole.

**Methods:** We first investigated the relationship between *Plasmodium falciparum* malaria and NTS bacteremia in all pediatric admissions aged ≤5 years between August 1998 and October 2019 (n=75,015). We then assayed hepcidin and measures of iron status in five groups: (1) children with concomitant severe malaria anemia (SMA) and NTS (SMA+NTS, n=16); and in matched children with (2) SMA alone (n=33); (3) NTS alone (n=33); (4) cerebral malaria (CM, n=34); and (5) community-based children.

**Results:** In hospitalized children SMA, but not other malaria phenotypes, was associated with an increased risk of NTS (adjusted OR 2.88 [95% CI 1.97, 4.23]; P<0.0001). Risk of NTS increased by 30% with each 1g/dl decrease in hemoglobin concentrations. In hospitalized children median hepcidin levels were lower in the SMA+NTS (9.3 ng/mL [interquartile range 4.7, 49.8]) and SMA (31.1 ng/mL [5.5, 61.2]) groups, compared to levels in those with CM (90.7 ng/mL [38.7, 176.1]) or NTS (105.8 ng/mL [17.3, 233.3]), despite similar ferritin and CRP levels. Soluble transferrin receptor levels were lower in the CM group compared to the other hospitalized groups.

**Conclusion:** SMA was associated with increased risk of NTS and with reduced hepcidin levels. We hypothesized that reduced hepcidin might allow increased movement of iron into the *Salmonella*-containing vacuole favoring bacterial growth.

## Introduction

Malaria and invasive non-typhoidal *Salmonella* (NTS) are major causes of illness and death among children living in sub-Saharan Africa.According to the World Health Organization (WHO), 94% of the 409,000 malaria-associated deaths in 2019 occurred in the sub-Saharan African region, with children under five years of age being disproportionately vulnerable [1]. In this region, NTS bacteremia is also common accounting for 80% of the estimated 535,000 global cases in 2017 [2]. NTS bacteremia is associated with life-threatening sepsis in African children with case fatality rates of 20-25% [2, 3] and is highly prevalent in areas with concurrent malaria endemicity [4, 5].

NTS bacteremia and malaria infections are associated, and reduced malaria incidence has been associated with decreases in NTS cases [6, 7]. Epidemiological studies report positive associations between malaria, severe malarial anemia (SMA) and NTS bacteremia [8, 9], although these associations have not been observed universally [10, 11]. Studies report a number of contributory pathways by which malaria may increase susceptibility to NTS bacteremia including sustained hemolysis, accumulation of free heme from lysed red blood cells, increased gut permeability and upregulation of heme oxygenase-1 ([12], Figure 1).

**Figure 1.**
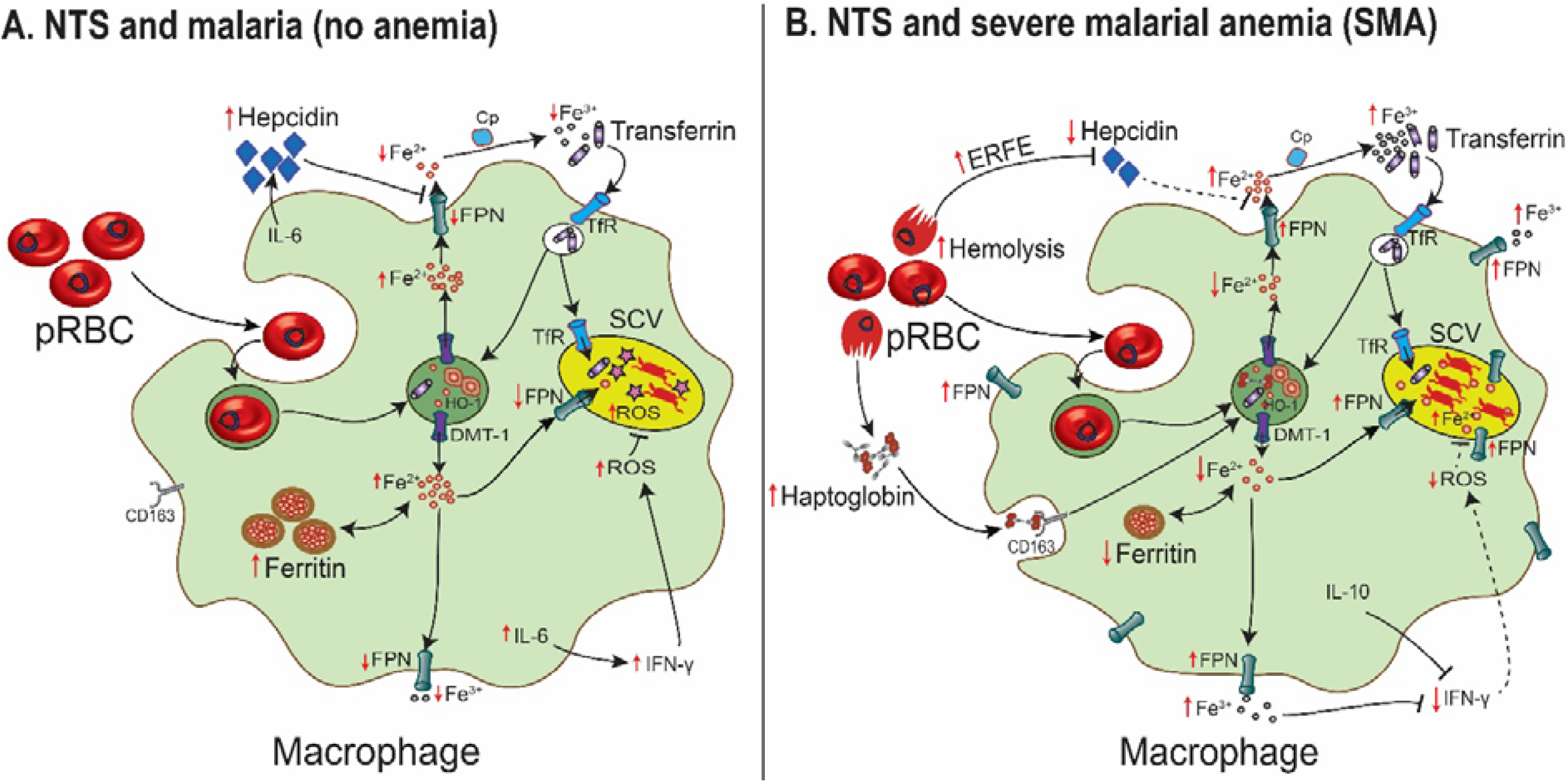
Low hepcidin levels in children with severe malarial anemia (SMA) may contribute to risk of non-typhoidal *Salmonella* (NTS) bacteremia. A) During malaria infections, proinflammatory responses and parasitemia induce expression of hepcidin, which sequesters iron into storage proteins including ferritin. Hepcidin also degrades ferroportin (FPN) on the macrophage membrane and *Salmonella* containing vacuole (SCV) [18], resulting in decreased iron availability for NTS bacteria. However, the bacteria may utilize other iron acquisition strategies including recruiting transferrin receptors (TfR) in early endosomes. Proinflammatory responses, including production of interleukin (IL)-6, the main hepcidin inducer, and interferon-gamma (IFN-γ) mediate killing of NTS through induction of reactive oxygen species (ROS) and other pathways. B) In SMA, increased hemolysis and erythropoietic drive induce production of erythroferrone (ERFE) [35], a hormone that downregulates the synthesis of hepcidin. As a result, there is increased expression of ferroportin (FPN) on the surface of the macrophage and the SCV [18]. Heme from hemolyzed parasitized red blood cells (pRBC) and the haptoglobin-hemoglobin complex is broken down by heme oxygenase-1 (HO-1) into equimolar amounts of iron, biliverdin and carbon monoxide. HO-1 and its products (bilirubin and carbon-monoxide) downregulate immune responses against NTS [13]. The net effect of low hepcidin, HO-1, SMA-induced cytokines such as IL-10, and increased intracellular iron levels is increased flow of iron into the SCV through FPN and reduced immune clearance of the bacteria. DMT-1 denotes divalent metal transporter 1; and Cp, ceruloplasmin. Red arrows indicate direction of increase or decrease; dotted lines indicate reduced activity.

Heme oxygenase-1 counters the toxic effects of free heme, impairs neutrophil oxidative burst capacity, reduces neutrophil bactericidal activity, and promotes iron accumulation in macrophages [13, 14]. *In vitro* and animal studies suggest that hepcidin, the master iron regulator, may also play an important role by controlling the availability of iron [15–17], a nutrient critical for NTS growth and proliferation [14, 16]. Hepcidin degrades ferroportin, the sole iron exporter, which was recently shown to transport iron into the *Salmonella*-containing vacuole (SCV) [18, 19]. In murine studies, low hepcidin levels and increased ferroportin expression on the SCV are associated with increased susceptibility to *Salmonella* Typhimurium infections [15, 19] (Figure 1). However, there are no studies of hepcidin in NTS infection in humans. In this study, we investigated the relationship between malaria and NTS in 75,015 hospitalized Kenyan children over a 21-year period and then estimated levels of hepcidin and other iron biomarkers in children with NTS bacteremia and malaria.

## Methods

### Study design and participants

The study was conducted in Kilifi, a rural malaria-endemic area along the Kenyan coast. The estimated incidence rate of NTS bacteremia among children ≤5 years was 36.6 cases/100,000 person-years between 1998 t0 2014 [20]. Pediatric admissions to Kilifi County Hospital have had routine clinical and laboratory data, including complete blood count, malaria microscopy, blood cultures and a stored plasma sample collected since August 1998. Our study included: a study of all pediatric admissions over a 21-year period and 2) a sub-study assessing iron status and hepcidin levels in subgroups of children with various malaria phenotypes and NTS.

#### 1) Study of all pediatric admissions

To confirm previous reports of associations between malaria and NTS, we first investigated the relationship between malaria and NTS bacteremia among hospitalized children with complete age and hemoglobin data (Figure 2). We analyzed retrospective data from 75,015 pediatric admissions aged ≤60 months between 1^st^ August 1998 and 31^st^ October 2019.

**Figure 2.**
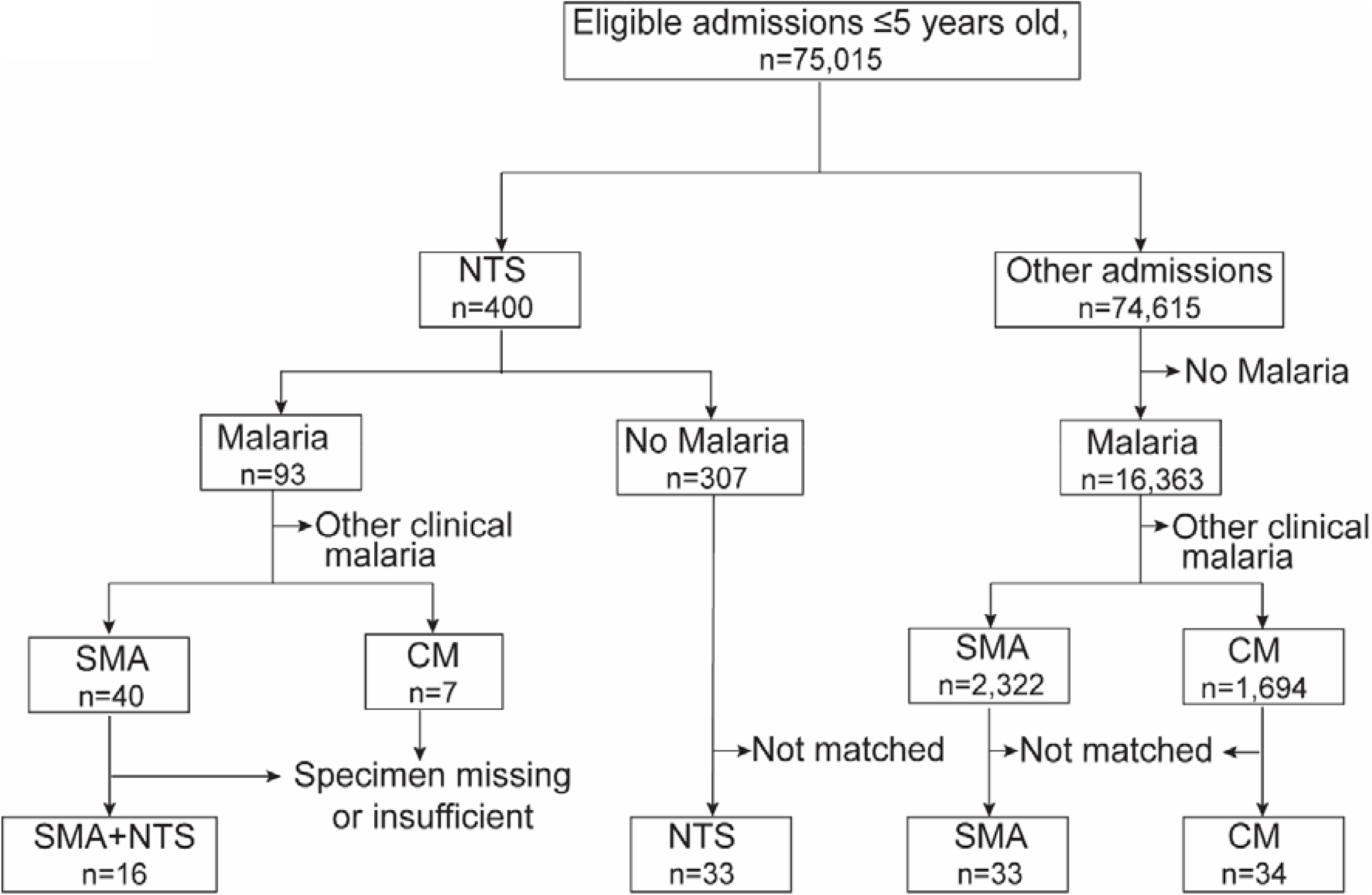
Selection of study participants. All children aged ≤60 months with complete age and hemoglobin data admitted between August 1998 and October 2019 were included in the retrospective epidemiological analysis. Children with concomitant severe malaria and non-typhoidal *Salmonella* (NTS), and whose specimens were available in the Kilifi biobank, were enrolled into the iron and hepcidin sub-study. Each child was then matched with two hospitalized children with NTS alone, severe malaria anemia (SMA) alone, and cerebral malaria (CM) alone based on age, sex and year of admission.

#### 2) Hepcidin sub-study

We then assessed for iron status and inflammation using stored samples from five groups of children including those hospitalized with: 1) SMA and NTS coinfection (SMA+NTS); 2) SMA alone; 3) NTS alone; and 4) cerebral malaria (CM) and 5) community-based children with and without asymptomatic malaria. A total of 40 children had SMA+NTS, and sufficient volumes of stored plasma for iron and inflammatory biomarker analyses were available for 16 children. Few children had CM and NTS coinfection (n=7) and samples were either missing or insufficient for analysis. Each child from group 1 was then matched with two from each of the other hospitalized groups based on age, sex and year of admission (Figure 2).

Community-based children were from the Ngerenya cohort, an ongoing, rolling study evaluating malaria immunity, as previously described [21, 22]. Within this cohort, children are followed to a maximum age of 13 years with annual cross-sectional bleeds. Iron biomarkers were measured from a single cross-sectional bleed based on the availability of plasma samples archived at −80°C.

### Laboratory procedures

Blood was examined for malaria parasites using Giemsa-stained thick and thin blood films and complete blood count (Beckman Coulter) was measured. Samples for bacterial culture were collected in BACTEC^®^ Peds Plus bottles and processed with a BACTEC 8050 automated blood-culture instrument (Becton-Dickson, UK). Positive samples were sub-cultured [23] and serological tests and biochemical test kits (API, bioMérieux) were used to confirm suspected pathogens. The following were considered to be contaminants: *Bacillus* species, *Micrococcus* species, viridans group *Streptococcus*, coagulase negative *Staphylococcus*, and coryneforms. We assayed hepcidin (Hepcidin-25 [human] EIA kit; Bachem), soluble transferrin receptor (sTfR; enzyme-linked immunosorbent assay; R&D systems), ferritin (micro-particle enzyme immunoassay, IMx [MEIA] ferritin assay, Abbott Laboratories), and C-reactive protein (CRP, Dade Dimension particle enhanced turbidimetric immunoassay; Hitachi Corp.). Hepcidin/ferritin ratio was calculated by dividing hepcidin (ng/ml) by ferritin (µg/L). Rapid antibody tests were used to perform systematic HIV-1 testing according to national policies from 2007. Sickle cell disease was diagnosed clinically.

### Clinical definitions

In classifying children with *Plasmodium falciparum* malaria, we defined any malaria as any hospitalized child with malaria parasitemia; SMA as hemoglobin <5 g/dl; CM as Blantyre coma score <3 according to WHO criteria [24], and asymptomatic malaria as those without clinical features. Severe anemia in children without malaria was defined as hemoglobin <5 g/dl. NTS bacteremia was defined as isolation of *Salmonella enterica* subspecies excluding *Salmonella enterica* Typhi or Paratyphi in blood cultures. Stunting was defined as height-for-age z-score <–2; wasting as weight-for-height z-score <–2 or mid-upper arm circumference <12.5 cm in children >6 months of age; and underweight as weight-for-age z-score < –2 using WHO Child Growth Standards [25].

### Statistical analysis

All data were analyzed using STATA 15.1 for Windows (StatCorp. College Station, Texas, USA). In the study of all pediatric admissions, we reported categorical data in numbers and corresponding percentages. We used univariable and multivariable logistic regression models (adjusted for age, sex, and underweight) to investigate for putative risk factors for NTS bacteremia in hospital admissions. We used a causal directed acyclic graph, as previously described [26], to assess the suitability of covariates for use in the multivariable model (Supplementary Figure 1). Variables with P values of < 0.1 in the univariable analysis were included in a multivariate model. In the hepcidin sub-study, continuous data were reported as medians and interquartile ranges (IQR) and compared using the Wilcoxon rank-sum test. We normalised nonnormally distributed variables by log_e_-transformation, and then analysed using multivariable linear regression models to adjust for potential confounders. Pearson’s correlation was also used as appropriate.

### Ethics

Ethical approval was granted by the Scientific Ethics Review Unit of the Kenya Medical Research Institute. Individual written informed consent was provided by parents or guardians of study participants.

## Results

### Study of all hospital admissions

A total of 75,015 children aged ≤60 months were admitted to Kilifi County Hospital during the 21-year study period and had complete data for analysis. Median age was 11.8 months (IQR 2.2, 26.1) and 42,444 (56.6%) were male. *P. falciparum* malaria was identified in the blood films of 16,456 (21.9%) hospitalized children; of whom 2,362 (14.4%) had SMA, 1,701 (10.3%) had CM, and 343 (2.1%) had concomitant SMA and CM. Pathogenic bacterial organisms were isolated from 3,846 (5.1%) blood cultures. NTS bacteremia was identified in 400 (10.4%) of the positive blood cultures. Of the NTS isolates, 306 were serotyped and 45.1% (138/306) were *Salmonella enterica* serovar Enteritidis and 44.4% (136/306) were serovar Typhimurium, while 10.7% (32/306) were not typeable.

NTS bacteremia was identified in 93/16,456 (0.6%) hospitalized children with *P. falciparum* malaria, including 40/2,362 (1.7%) with SMA and 7/1,701 (0.4%) with CM. SMA was associated with an almost three-fold increased risk of NTS bacteremia (adj. OR 2.88 [95% CI 1.97, 4.23]; adj. P<0.0001). However, malaria parasitemia (adj. OR 1.18 [95% CI 0.90, 1.54]; P=0.23), CM (adj. OR 1.11 [95% CI 0.51, 2.39]; adj. P=0.80) and other malaria phenotypes excluding SMA (adj. OR 0.77 [95% CI 0.55, 1.07; adj. P=0.12) were not associated with increased risk of NTS bacteremia in models adjusted for age, sex and underweight (Table 1). An increase of 1 g/dl in hemoglobin levels was associated with a 23% reduction in the risk of NTS bacteremia in all children (adj. OR 0.77 [95% CI 0.74, 0.80]; P<0.0001, Table 1); and a 30% reduction in the risk of NTS bacteremia in children with malaria (adj. OR 0.70 [95% CI 0.63, 0.77]; adj. P<0.0001). Hemoglobin levels remained a significant predictor of the risk of NTS bacteremia in a multivariate model (Supplementary Table 1). Fever, diarrhea, very severe pneumonia, wasting, underweight, HIV status, and sickle cell disease were also associated with NTS bacteremia as shown in Table 1.

**Table 1.** Factors associated with non-typhoidal *Salmonella* bacteremia in all hospitalized children (n=75,015) between August 1998 and October 2019

| Characteristic | NTS, n (%) | Hospital controls, n (%) | OR (95% CI) | P | Adj. OR (95% CI)<sup>a</sup> | Adj. P<sup>a</sup> |
| --- | --- | --- | --- | --- | --- | --- |
| **Clinical features** |  |  |  |  |  |  |
| Age, years | 400 | 74,615 | 1.00 (0.94, 1.08) | 0.87 | 0.98 (0.91, 1.07) | 0.68 |
| Sex, male | 224/400 (56.0) | 42,220/74,614 (56.6) | 0.98 (0.80, 1.19) | 0.81 | 0.96 (0.77, 1.19) | 0.69 |
| Fever (temperature >37.5°C) | 267/369 (72.4) | 37,886/61,963 (61.1) | 1.66 (1.32, 2.09) | <0.0001 | 1.94 (1.50, 2.51) | <0.0001 |
| Diarrhea<sup>b</sup> | 126/400 (31.5) | 14,185/74,596 (19.0) | 1.96 (1.58, 2.42) | <0.0001 | 1.67 (1.32, 2.11) | <0.0001 |
| Vomiting<sup>c</sup> | 127/388 (32.7) | 18,055/73,223 (24.7) | 1.48 (1.20, 1.84) | <0.0001 | 1.34 (1.06, 1.69) | 0.02 |
| Severe pneumonia<sup>d</sup> | 111/400 (27.8) | 21,063/74,591 (28.2) | 0.98 (0.78, 1.22) | 0.83 | 0.97 (0.76, 1.23) | 0.77 |
| Very severe pneumonia<sup>e</sup> | 59/400 (14.8) | 7480/74,587 (10.0) | 1.55 (1.17, 2.05) | 0.002 | 1.55 (1.15, 2.10) | 0.004 |
| Wasting | 180/380 (47.4) | 19,487/67,474 (28.9) | 2.22 (1.81, 2.71) | <0.0001 | 1.33 (1.02, 1.72) | 0.04 |
| Stunting | 198/371 (53.4) | 27,802/69,492 (40.0) | 1.72 (1.40, 2.11) | <0.0001 | 1.06 (0.82, 1.38) | 0.65 |
| Underweight | 204/340 (60.0) | 28,470/68,642 (41.5) | 2.12 (1.70, 2.63) | <0.0001 | 2.12 (1.70, 2.63) | <0.0001 |
| **Laboratory characteristics** |  |  |  |  |  |  |
| Hemoglobin, g/dl | 400 | 74,615 | 0.78 (0.76, 0.81) | <0.0001 | 0.77 (0.74, 0.80) | <0.0001 |
| Any malaria <sup>f</sup> | 93/400 (23.3) | 16,363/74,613 (21.9) | 1.08 (0.85, 1.36) | 0.53 | 1.18 (0.90, 1.54) | 0.23 |
| SMA <sup>g</sup> | 40/347 (11.5) | 2,322/60,572 (3.8) | 3.27 (2.34, 4.56) | <0.0001 | 2.88 (1.97, 4.22) | <0.0001 |
| CM <sup>g</sup> | 7/241 (2.9) | 1,694/54,420 (3.1) | 0.93 (0.44, 1.98) | 0.51 | 1.11 (0.51, 2.39) | 0.80 |
| Non-SMA malaria | 48/355 (13.5) | 13,703/71,953 (19.0%) | 0.66 (0.49, 0.90) | 0.009 | 0.77 (0.55, 1.07) | 0.12 |
| HIV status, positive <sup>h</sup> | 38/139 (27.3) | 1,756/35,327 (5.0) | 7.19 (4.94, 10.48) | <0.0001 | 5.43 (3.49, 8.47) | <0.0001 |
| Sickle cell disease | 14/400 (3.7) | 1,115/74,589 (1.5) | 2.39 (1.39, 4.09) | 0.001 | 2.02 (1.07, 3.84) | 0.03 |

Abbreviations: NTS, non-typhoidal *Salmonella*; OR, odds ratio; CI, confidence interval; SMA, severe malaria anemia; and CM, cerebral malaria; and HIV, human immunodeficiency virus. <sup>a</sup> Adjusted (adj.) OR and P values were derived from logistic regression models adjusting for age in years, sex, and underweight as appropriate; <sup>b</sup> Passage of three or more loose or liquid stools within 24 hours; <sup>c</sup> Oral eviction of gastrointestinal contents; <sup>d</sup> History of cough or difficulty in breathing plus lower chest wall indrawing; <sup>e</sup> Cough or difficulty breathing plus either prostration, lethargy, hypoxia, loss of consciousness, or a history of convulsions; <sup>f</sup> All hospitalized children with malaria parasitemia on blood film; <sup>g</sup> Children with overlapping SMA and CM clinical syndromes were excluded from analysis. <sup>h</sup> Data was available from February 2005 after routine HIV testing was introduced.

### Hepcidin sub-study

We included 116 hospitalized children in the following groups: 1) 16 with SMA+NTS; 2) 33 with SMA alone; 3) 33 with NTS alone; 4) 34 with CM; and also 5) community-based children with and without asymptomatic malaria parasitemia. The clinical characteristics of the children in the sub-study are shown in Supplementary Table 2.

### Hepcidin levels in children with malaria

We first compared hepcidin levels among children with malaria. Hepcidin levels were lower in children with SMA (median 31.1 ng/ml [IQR 5.5, 61.2]) compared to those with CM (90.7 ng/ml [IQR 38.7, 176.1]; P=0.002). However, both of these severe malaria groups had higher hepcidin levels than children with asymptomatic malaria parasitemia living in the community (Figure 3A). Children with asymptomatic parasitemia had marginally increased hepcidin levels (6.5 ng/ml [IQR 2.0, 13.1]) compared to those without parasitemia (3.8 ng/ml [IQR 1.2, 12.6]; P=0.13).

**Figure 3.**
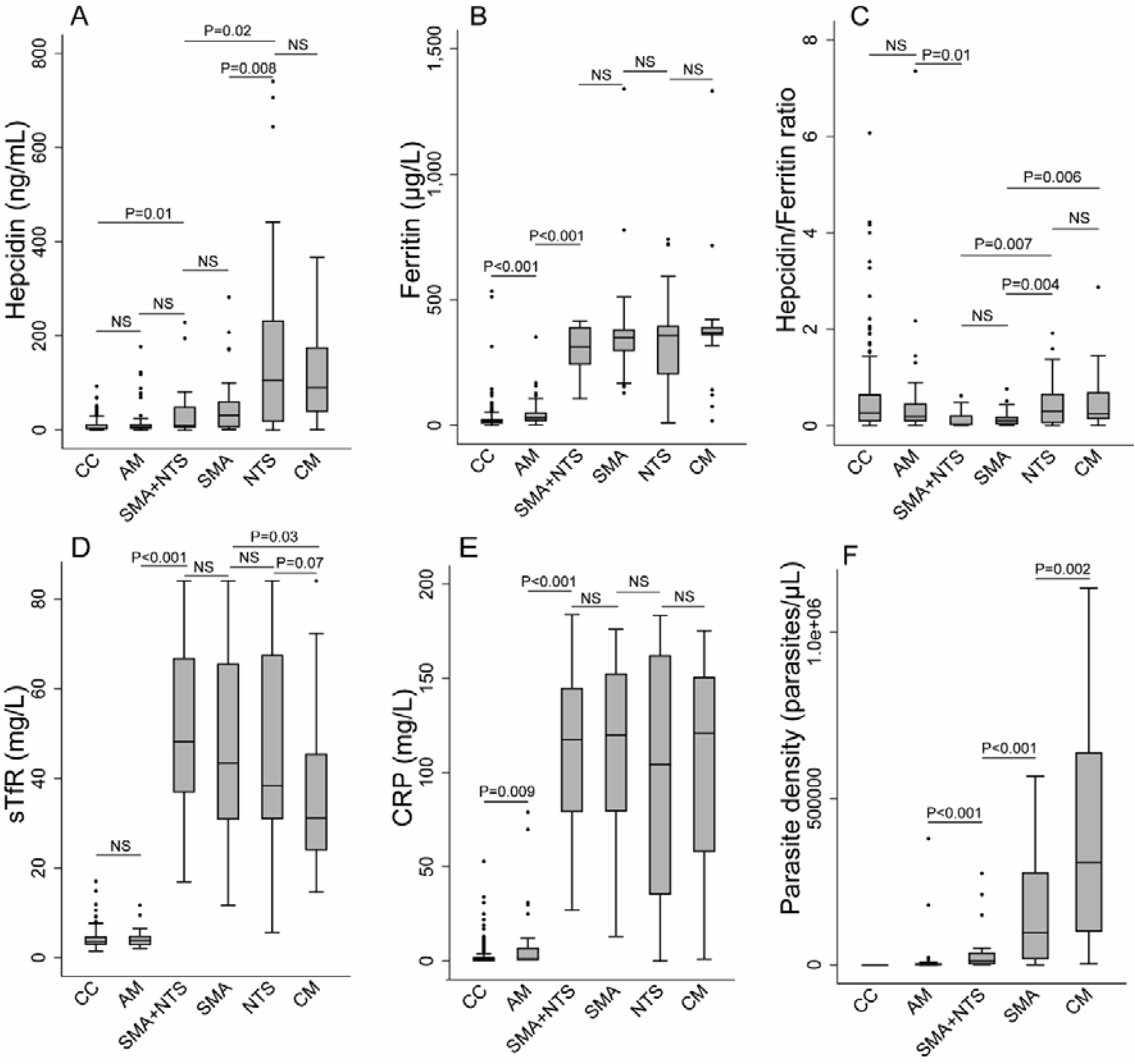
Circulating levels of (A) hepcidin (B) ferritin (C) hepcidin/ferritin ratio (D) soluble transferrin receptors (sTfR) (E) C-reactive protein (CRP) and (F) hemoglobin. P values were determined by the Wilcoxon rank-sum test. ‘NS’ indicates P > 0.05. CC denotes community-based children without malaria; AM, asymptomatic malaria; CM, cerebral malaria; SMA, severe malaria anemia; NTS, non-typhoidal Salmonella; SMA+NTS, SMA and NTS coinfection.

We then explored differences in putative regulators of hepcidin. Children with SMA had increased erythropoietic drive as indicated by higher sTfR levels (43.3 mg/L [IQR 30.8, 65.6]) than those with CM (31.2 mg/L [IQR 23.9, 45.5]; P=0.03), although ferritin and CRP levels did not differ between the groups (Figure 3). Children with SMA also had a lower hepcidin/ferritin ratio (0.10 [IQR 0.03, 0.19]) than those with CM (0.24 [0.14, 0.69]; P=0.006), or asymptomatic parasitemia (0.19 [0.09, 0.46]; P=0.01; Figure 3C) indicating that hepcidin levels were less likely to be explained by ferritin levels. Hospitalized children had higher levels of ferritin, sTfR, and CRP and higher parasite densities than those living in the community (Figure 3 B, D-F).

### Hepcidin levels in children with malaria and NTS

We then considered hepcidin levels in children with malaria and NTS. Hepcidin levels were lower in children with SMA+NTS (9.3 ng/ml [IQR 4.7, 49.8]) and SMA alone (31.1 ng/ml [IQR 5.5, 61.2]); P=0.43), compared to those with NTS alone (105.8 ng/ml [IQR 17.3, 233.3]; Figure 3A, Table 2). In a linear regression model controlled for age, sex and underweight hepcidin levels were 2-fold higher in children with NTS (adj. β 2.02 [95% CI 0.72, 3.31]; adj. P=0.002) compared to those with SMA+NTS (Supplementary Table 3), although sTfR, ferritin, and CRP levels did not differ between the groups (Figure 3 B, D, E). Hepcidin/ferritin ratios were lower in children with SMA+NTS (0.03 [IQR 0.01, 0.22]) compared to NTS alone (0.31 [IQR 0.06, 0.66]; P=0.007; Table 2, Figure 3C). *P. falciparum* parasite densities varied between groups with parasite densities decreasing between the CM, SMA, SMA+NTS and asymptomatic parasitemia groups (Figure 3F).

**Table 2.**
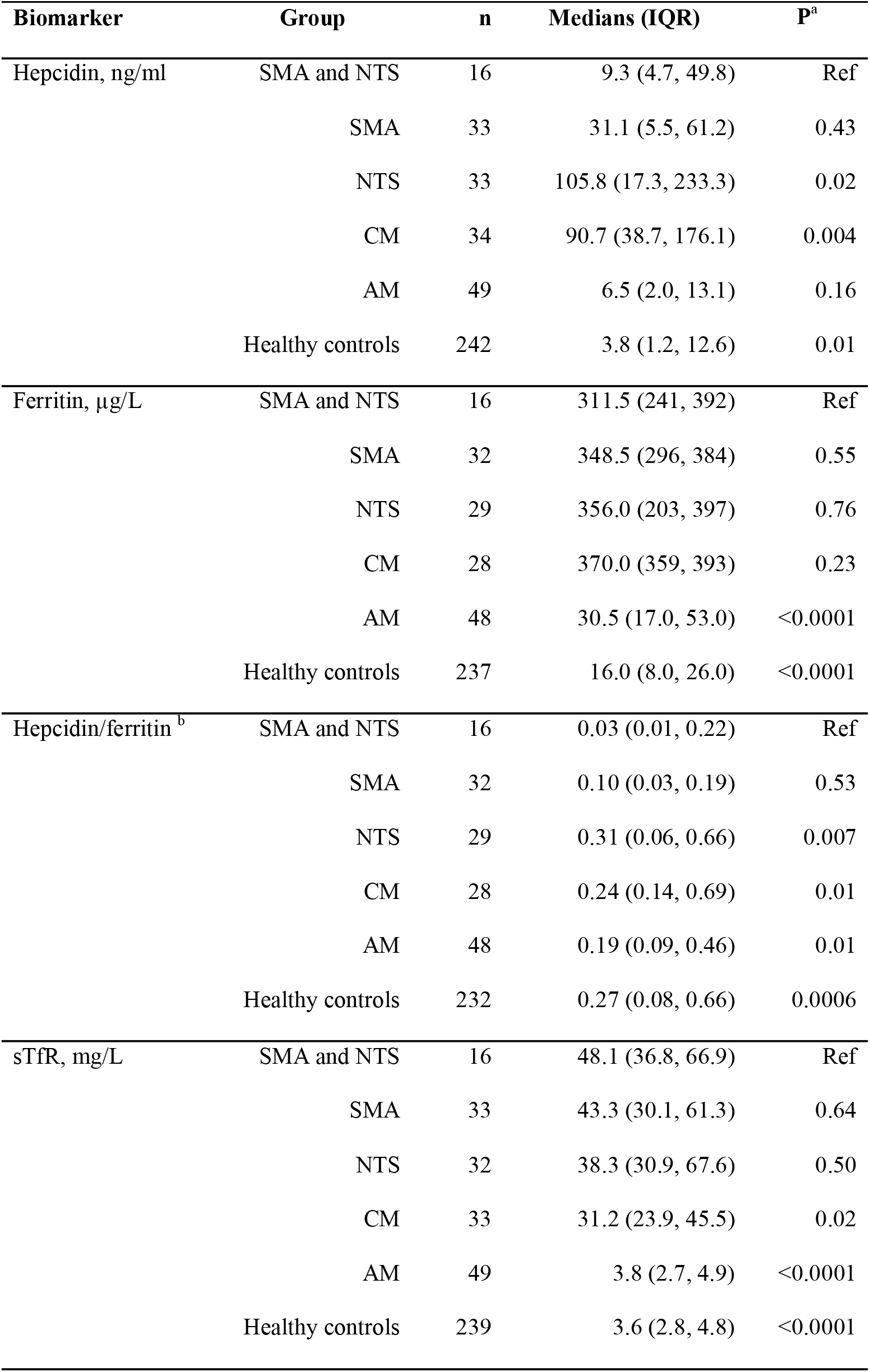
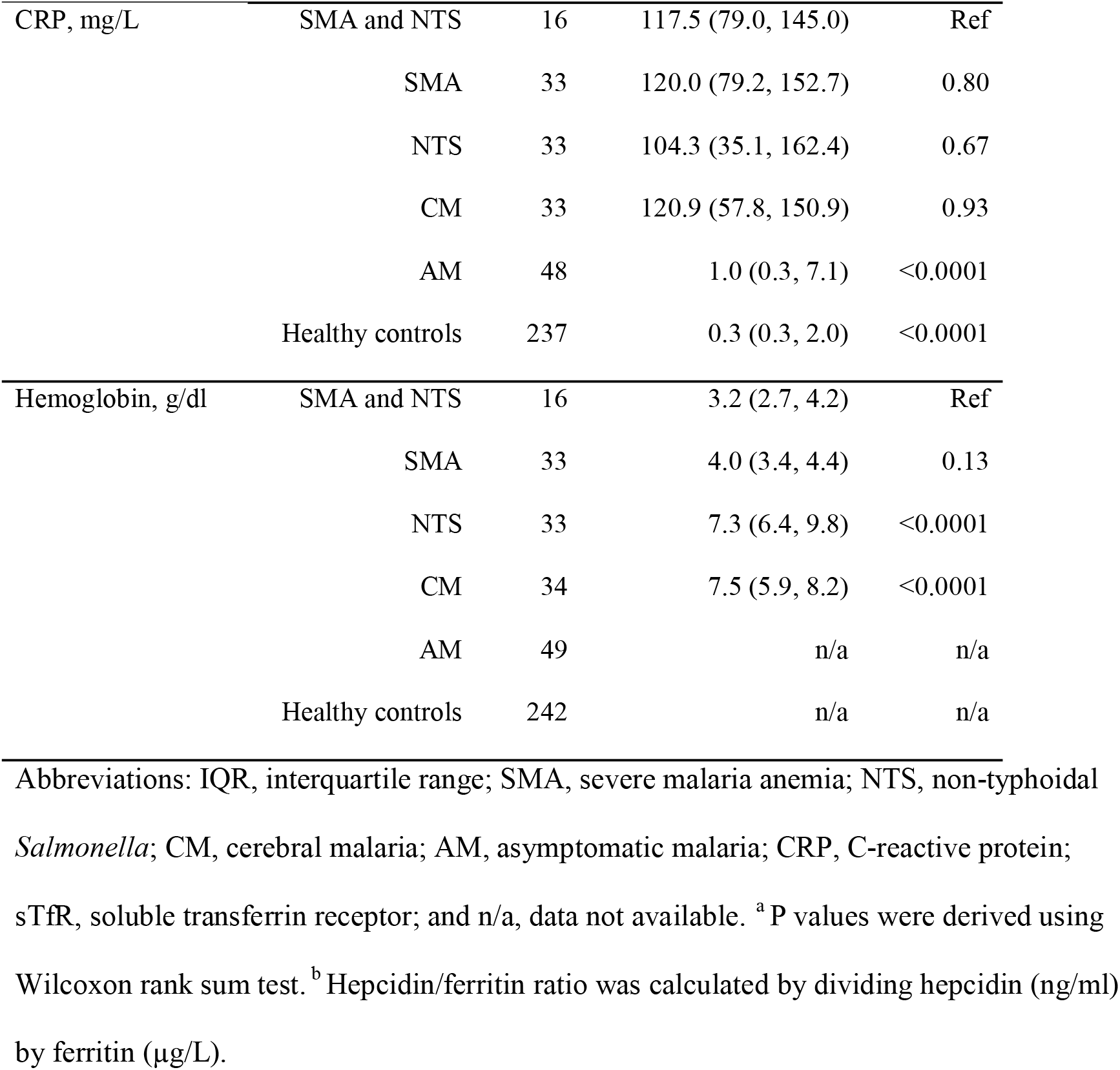
Hepcidin and biomarkers of iron status and inflammation in a sub-study of hospitalized and community-based children

| **Biomarker** | **Group** | **n** | **Medians (IQR)** | **P<sup>a</sup>** |
| --- | --- | --- | --- | --- |
| Hepcidin, ng/ml | SMA and NTS | 16 | 9.3 (4.7, 49.8) | Ref |
|  | SMA | 33 | 31.1 (5.5, 61.2) | 0.43 |
|  | NTS | 33 | 105.8 (17.3, 233.3) | 0.02 |
|  | CM | 34 | 90.7 (38.7, 176.1) | 0.004 |
|  | AM | 49 | 6.5 (2.0, 13.1) | 0.16 |
|  | Healthy controls | 242 | 3.8 (1.2, 12.6) | 0.01 |
| Ferritin, µg/L | SMA and NTS | 16 | 311.5 (241, 392) | Ref |
|  | SMA | 32 | 348.5 (296, 384) | 0.55 |
|  | NTS | 29 | 356.0 (203, 397) | 0.76 |
|  | CM | 28 | 370.0 (359, 393) | 0.23 |
|  | AM | 48 | 30.5 (17.0, 53.0) | <0.0001 |
|  | Healthy controls | 237 | 16.0 (8.0, 26.0) | <0.0001 |
| Hepcidin/ferritin <sup>b</sup> | SMA and NTS | 16 | 0.03 (0.01, 0.22) | Ref |
|  | SMA | 32 | 0.10 (0.03, 0.19) | 0.53 |
|  | NTS | 29 | 0.31 (0.06, 0.66) | 0.007 |
|  | CM | 28 | 0.24 (0.14, 0.69) | 0.01 |
|  | AM | 48 | 0.19 (0.09, 0.46) | 0.01 |
|  | Healthy controls | 232 | 0.27 (0.08, 0.66) | 0.0006 |
| sTfR, mg/L | SMA and NTS | 16 | 48.1 (36.8, 66.9) | Ref |
|  | SMA | 33 | 43.3 (30.1, 61.3) | 0.64 |
|  | NTS | 32 | 38.3 (30.9, 67.6) | 0.50 |
|  | CM | 33 | 31.2 (23.9, 45.5) | 0.02 |
|  | AM | 49 | 3.8 (2.7, 4.9) | <0.0001 |
|  | Healthy controls | 239 | 3.6 (2.8, 4.8) | <0.0001 |
| CRP, mg/L | SMA and NTS | 16 | 117.5 (79.0, 145.0) | Ref |
|  | SMA | 33 | 120.0 (79.2, 152.7) | 0.80 |
|  | NTS | 33 | 104.3 (35.1, 162.4) | 0.67 |
|  | CM | 33 | 120.9 (57.8, 150.9) | 0.93 |
|  | AM | 48 | 1.0 (0.3, 7.1) | <0.0001 |
|  | Healthy controls | 237 | 0.3 (0.3, 2.0) | <0.0001 |
| Hemoglobin, g/dl | SMA and NTS | 16 | 3.2 (2.7, 4.2) | Ref |
|  | SMA | 33 | 4.0 (3.4, 4.4) | 0.13 |
|  | NTS | 33 | 7.3 (6.4, 9.8) | <0.0001 |
|  | CM | 34 | 7.5 (5.9, 8.2) | <0.0001 |
|  | AM | 49 | n/a | n/a |
|  | Healthy controls | 242 | n/a | n/a |

Abbreviations: IQR, interquartile range; SMA, severe malaria anemia; NTS, non-typhoidal *Salmonella*; CM, cerebral malaria; AM, asymptomatic malaria; CRP, C-reactive protein; sTfR, soluble transferrin receptor; and n/a, data not available. <sup>a</sup> P values were derived using Wilcoxon rank sum test. <sup>b</sup> Hepcidin/ferritin ratio was calculated by dividing hepcidin (ng/ml) by ferritin (μg/L).

Log-hepcidin levels were positively correlated with log-ferritin (r=0.35, P=0.0002); log-CRP (r=0.32, P=0.0005); hemoglobin levels (r=0.25, P=0.008); and parasite density (r=0.44, P<0.0001) among the hospitalized children. We also found a negative correlation between log-hepcidin and log-sTfR (r= –0.36, P<0.0001). However, the direction and strength of correlation between log-hepcidin and its predictors varied across individual groups as shown in Supplementary Table 4.

## Discussion

Malaria and NTS are important causes of hospitalization and death among children living in sub-Saharan Africa [1, 2]. In this study, we analyzed retrospective data from 75,015 hospitalized children aged ≤60 months and found that SMA, but not CM or other malaria phenotypes, was associated with increased risk of NTS bacteremia. In a sub-study investigating iron biomarkers, children with SMA and SMA+NTS had lower hepcidin levels, than those with NTS or CM alone. We did not find differences in ferritin or CRP levels among the hospitalized children, but those with SMA+NTS, and SMA or NTS alone, had higher sTfR levels than those with CM. Children with asymptomatic parasitemia had lower levels of hepcidin, ferritin, CRP and sTfR and lower parasite densities than hospitalized children.

SMA was associated with a three-fold increased risk of NTS bacteremia among hospitalized children. However, hospitalized children with malaria parasitemia, CM, or other malaria phenotypes without severe anemia, did not have an increased risk of NTS bacteremia. Moreover, each 1g/dl increase in hemoglobin concentrations in children with malaria was associated with a 30% reduction in risk of NTS bacteremia. Previous studies across sub-Saharan Africa have also reported an increased risk of NTS bacteremia in children with SMA [8, 9], but not CM [9, 27]. A study in Malawian children with severe malaria reported a 43% increase in the risk of NTS bacteremia per 1g/dl reduction in hemoglobin levels [8]. In contrast, a study in Mozambican children reported no clear-cut association between SMA and NTS bacteremia, although few children had NTS bacteremia (n=12) [10]. In agreement with the current study, previous studies found no association between any malaria parasitemia and risk of NTS bacteremia [4, 20], although other studies have reported mixed findings with malaria both reducing [28, 29] and increasing [7, 30] risk of NTS bacteremia. These differences might be explained by the prevalence of malarial anemia within the study populations or various other factors, including nutritional status. Taken together, our findings suggest that SMA, rather than other malarial phenotypes, underlies the association between malaria and NTS bacteremia.

A number of pathways may contribute to increased risk of NTS bacteremia in children with SMA including hemolysis, iron overload and upregulation of heme oxygenase-1 [12] (Figure 1). Hepcidin may also influence risk of NTS bacteremia in SMA by controlling the availability of iron for bacterial growth [15–17]. Hepcidin levels differed between the various malaria phenotypes. We observed that hepcidin levels were lower in children with SMA compared to those with CM in agreement with a study in Kenyan children, which found lower hepcidin levels in malaria patients with severe anemia compared to those with higher hemoglobin levels [31]. In contrast, a study in Nigerian children found no difference in hepcidin levels between children with SMA and CM and higher hepcidin levels in uncomplicated compared to severe malaria [32]. In agreement with previous studies [33, 34], we found higher hepcidin levels in children with severe malaria compared to those with asymptomatic parasitemia. Our findings may be explained by higher sTfR levels in SMA compared to CM, indicating increased erythropoietic activity. Severe anemia negatively regulates hepcidin production through the action of erythroferrone [35], even in the presence of inflammation or infection [36, 37]. Inflammation, as measured by ferritin and CRP, did not differ between the SMA and CM groups, although parasite density, known to correlate with hepcidin levels [38], was higher in CM. Lower levels of hepcidin in children with asymptomatic parasitemia might be explained by reduced inflammation.

Little is known about hepcidin during NTS and malaria infection in humans, although murine studies indicate that reduced hepcidin levels increase susceptibility to *Salmonella* infection during hemolysis [15]. In the current study, children with SMA+NTS and SMA had lower hepcidin levels than those with NTS alone; although sTFR, CRP and ferritin levels did not differ between these groups. A challenge infection study with *Salmonella* Typhi in the United Kingdom found higher hepcidin concentrations during acute infection [39]. *In-vitro* and murine studies show that NTS may directly or indirectly upregulate hepcidin expression and perturb iron regulatory pathways [17]. Recent evidence showed that hepcidin degrades ferroportin on the *Salmonella* containing vacuole (SCV) thus limiting iron movement into the SCV [18]. However, whether iron accumulation in the SCV promotes bacterial growth [16, 19] or kills bacteria through the Fenton reaction [18] remains controversial. Low hepcidin levels in mice with severe hemolytic anemia were associated with increased susceptibility to *Salmonella* Typhimurium infection and hepcidin treatment improved survival [15]. We speculate that low hepcidin levels in children with SMA and SMA+NTS might contribute to increased iron transport through ferroportin on the SCV, increasing iron availability for NTS growth (Figure 1). Surprisingly, sTfR levels were elevated in NTS alone despite higher hemoglobin levels. It is not known whether NTS might induce transcription of transferrin receptors to increase transferrin iron acquisition, although transferrin receptors were observed on the SCV during early phases of endocytosis in murine models [40].

To the best of our knowledge, this is the first study reporting hepcidin levels in children with NTS or with concomitant SMA and NTS bacteremia. Strengths of the study are that we utilized a very large 21-year dataset (n=75,015) with matching stored samples to identify and describe associations between severe malaria, NTS bacteremia and hepcidin. Our study has some important limitations. First, the study was observational, and as such associations may be subject to unmeasured confounders, reverse causality and collider bias. Second, we did not measure additional parameters such as serum iron, transferrin saturation, and haptoglobin levels due to volumes and availability of stored samples. Additionally, a few participants had sTfR concentrations above the cut-off values making it challenging to interpret findings from regression models for sTfR (Supplementary Table 3). Finally, our study was conducted in a single site and only 16 children with SMA+NTS bacteremia had samples with adequate volumes for testing. It is also possible that our study underestimated associations, considering the low sensitivity of blood cultures used to identify NTS. Nonetheless, this study complements the existing *in vitro* and animal data on the relationship between SMA and NTS bacteremia and provides preliminary evidence on the possible role of hepcidin in mediating this association.

In conclusion, SMA was associated with an increased risk of NTS bacteremia in children and reduced hepcidin levels were observed in children with SMA and SMA+NTS. Further studies are needed to understand the role of the hepcidin-ferroportin axis in susceptibility to NTS in human subjects, how hepcidin and iron disturbances might mediate susceptibility to bacteremia due to NTS or other organisms, and how *P. falciparum*, iron deficiency or other etiologies of severe anemia influence this relationship.

## Data Availability

Data are available under the terms of the Creative Commons Attribution 4.0 International license (CC-BY 4.0)

## Acknowledgments

The authors would like to thank the children who participated in this study and their parents/guardians. We also acknowledge Esther Muthumbi for her help in acquiring the NTS serotyping data. This manuscript was submitted for publication with the permission of the Director of the Kenya Medical Research Institute (KEMRI).

