## Supplementary for "Hepcidin regulation in Kenyan children with severe malaria and non-typhoidal *Salmonella* bacteremia"

**Supplementary Table 1.** Multivariate logistic regression of factors associated with non-typhoidal *Salmonella* bacteremia in all hospitalized children (n=75,015)

| Characteristic | NTS, n (%) | Hospital controls, n (%) | OR (95% CI) | P | Adj. OR (95% CI) [a] | Adj. P [a] |
|---|---|---|---|---|---|---|
| **Clinical features** | | | | | | |
| Age, years | 400 | 74,615 | 1.00 (0.94, 1.08) | 0.87 | | |
| Sex, male | 224/400 (56.0) | 42,220/74,614 (56.6) | 0.98 (0.80, 1.19) | 0.81 | | |
| Fever (temperature >37.5ºC) | 267/369 (72.4) | 37,886/61,963 (61.1) | 1.66 (1.32, 2.09) | <0.0001 | 1.67 (1.26, 2.22) | <0.0001 |
| Diarrhea | 126/400 (31.5) | 14,185/74,596 (19.0) | 1.96 (1.58, 2.42) | <0.0001 | 1.81 (1.20, 2.73) | 0.005 |
| Vomiting | 127/388 (32.7) | 18,055/73,223 (24.7) | 1.48 (1.20, 1.84) | <0.0001 | 0.87 (0.65, 1.15) | 0.32 |
| Severe pneumonia | 111/400 (27.8) | 21,063/74,591 (28.2) | 0.98 (0.78, 1.22) | 0.83 | | |
| Very severe pneumonia | 59/400 (14.8) | 7480/74,587 (10.0) | 1.55 (1.17, 2.05) | 0.002 | 1.50 (1.04, 2.16) | 0.03 |
| Wasting | 180/380 (47.4) | 19,487/67,474 (28.9) | 2.22 (1.81, 2.71) | <0.0001 | 1.45 (1.05, 1.99) | 0.02 |
| Stunting | 198/371 (53.4) | 27,802/69,492 (40.0) | 1.72 (1.40, 2.11) | <0.0001 | 1.29 (0.95, 1.75) | 0.11 |
| Underweight | 204/340 (60.0) | 28,470/68,642 (41.5) | 2.12 (1.70, 2.63) | <0.0001 | 1.41 (0.97, 2.05) | 0.07 |
| **Laboratory characteristics** | | | | | | |

| | | | | | | |
|---|---|---|---|---|---|---|
| Hemoglobin, g/dl | 400 | 74,615 | 0.78 (0.76, 0.81) | <0.0001 | 0.77 (0.73, 0.81) | <0.0001 |
| Any malaria parasitemia | 93/400 (23.3) | 16,363/74,613 (21.9) | 1.08 (0.85, 1.36) | 0.53 | | |
| SMA | 40/347 (11.5) | 2,322/60,572 (3.8) | 3.27 (2.34, 4.56) | <0.0001 | 0.69 (0.43, 1.10) | 0.12 |
| CM | 7/241 (2.9) | 1,694/54,420 (3.1) | 0.93 (0.44, 1.98) | 0.51 | | |
| Non-SMA malaria | 48/355 (13.5) | 13,703/71,953 (19.0%) | 0.66 (0.49, 0.90) | 0.009 | | |
| HIV status, Positive [b] | 38/139 (27.3) | 1,756/35,327 (5.0) | 7.19 (4.94, 10.48) | <0.0001 | | |
| Sickle cell disease | 14/400 (3.7) | 1,115/74,589 (1.5) | 2.39 (1.39, 4.09) | 0.001 | 1.08 (0.54, 2.15) | 0.83 |

Abbreviations: NTS, non-typhoidal *Salmonella;* OR, odds ratio; CI, confidence interval; HIV, human immunodeficiency virus; SMA, severe malaria anemia; and CM, cerebral malaria. [a] All factors with P<0.1, age and sex were included in the model. [b] HIV status was not included in the model due to small numbers (data for this variable was only available for children admitted between 2005-2019).

**Supplementary Table 2.** Demographic and clinical characteristics of children in sub-study with measurements of iron and inflammation

| **Characteristic**[*] | **Hospitalized children** | | | | **Community children** | |
|---|---|---|---|---|---|---|
| | SMA and NTS (%) | SMA (%) | NTS (%) | CM (%) | AM (%) | Healthy controls (%) |
| Age, months (IQR) [a] | 23.6 (11.4, 31.6) | 22.3 (16.6, 30.3) | 17.2 (5.5, 28.5) | 23.0 (13.6, 36.4) | 63.2 (40.1, 77.4) | 41.6 (21.7, 64.2) |
| Sex, male | 8/16 (50.0) | 15/33 (45.5) | 20/33 (60.6) | 13/34 (38.2) | 27/49 (55.1) | 134/242 (55.4) |
| Fever [b] | 7/16 (43.8) | 21/33 (63.6) | 18/32 (56.3) | 25/34 (73.5) | 3/21 (14.3) | 5/88 (5.7) |
| Vomiting | 7/16 (43.8) | 16/33 (18.2) | 9/33 (27.2) | 10/33 (30.3) | n/a | n/a |
| Wasting | 9/16 (56.3) | 8/33 (28.2) | 15/32 (45.5) | 8/32 (23.5) | n/a | n/a |
| Stunting | 9/16 (56.3) | 13/30 (43.3) | 14/29 (48.3) | 14/31 (45.2) | n/a | n/a |
| Underweight | 7/12 (58.3) | 11/32 (34.3) | 20/29 (69.0) | 16/34 (47.1) | n/a | n/a |
| Pallor [c] | 5/5 (100) | 24/25 (96.0) | 10/27 (37.0) | 20/33 (60.6) | n/a | n/a |
| Coma (BCS <3) | 0/3 (0) | 5/24 (20.8) | 1/22 (4.6) | 34/34 (100.0) | n/a | n/a |
| HIV status, positive [d] | 0/1 (0) | 2/9 (22.2) | 2/9 (18.2) | 0/17 (0) | n/a | n/a |
| Transfused | 14/16 (87.5) | 19/33 (57.6) | 3/33 (9.1) | 9/34 (26.5) | n/a | n/a |
| In-hospital mortality | 4/16 (25.0) | 3/33 (9.1) | 8/33 (24.2) | 4/34 (11.8) | n/a | n/a |

Abbreviations: SMA, severe malaria anemia; NTS, non-typhoidal *Salmonella*; CM, cerebral malaria; AM, asymptomatic malaria; BCS, Blantyre coma score; and HIV, human immunodeficiency virus. [*]Only age, gender and axillary temperature data were available for community children. [a] Medians and interquartile ranges (IQR) are presented. [b] Temperature >37.5ºC. [c] Pallor was defined clinically. [d] HIV data was only available for children admitted between 2005-2019.

**Supplementary Table 3.** Geometric means and linear regression analyses of iron and/or inflammatory biomarkers by hospital groups.

| **Biomarker** | **Group** | **n** | Geometric means (95% CI) | Univariable regression | | Multivariable regression | |
|---|---|---|---|---|---|---|---|
| | | | | β (95% CI) | P | Adj. β (95% CI) ¶ | Adj. P¶ |
| Log-hepcidin, ng/ml | SMA and NTS | 16 | 11.7 (4.0, 34.5) | Reference | | Reference | |
| | SMA | 33 | 21.4 (12.4, 36.8) | 0.60 (-0.55, 1.75) | 0.30 | 0.91 (-0.37, 2.19) | 0.16 |
| | NTS | 33 | 48.7 (20.3, 116.4) | 1.42 (0.28, 2.57) | 0.02 | 2.02 (0.72, 3.31) | 0.002 |
| | CM | 34 | 62.9 (37.2, 106.4) | 1.68 (0.54, 2.82) | 0.004 | 1.94 (0.67, 3.20) | 0.003 |
| Log-ferritin, µg/L | SMA and NTS | 16 | 287.0 (233.4, 352.9) | Reference | | Reference | |
| | SMA | 32 | 329.3 (280.7, 386.4) | 0.14 (-0.27, 0.54) | 0.50 | 0.06 (-0.35, 0.47) | 0.77 |
| | NTS | 29 | 268.6 (193.8, 372.3) | -0.07 (-0.48, 0.35) | 0.75 | 0.01 (-0.41, 0.44) | 0.95 |
| | CM | 28 | 305.5 (226.8, 411.5) | 0.06 (-0.35, 0.48) | 0.77 | -0.01 (-0.42, 0.41) | 0.98 |
| Log -hepcidin/ferritin | SMA and NTS | 16 | 0.04 (0.01, 0.12) | Reference | | Reference | |
| | SMA | 32 | 0.07 (0.04, 0.12) | 0.48 (-0.62, 1.59) | 0.39 | 0.85 (-0.42, 2.12) | 0.19 |
| | NTS | 29 | 0.18 (0.08, 0.40) | 1.48 (0.36, 2.60) | 0.01 | 2.00 (0.70, 3.30) | 0.003 |
| | CM | 28 | 0.20 (0.11, 0.38) | 1.60 (0.47, 2.73) | 0.006 | 1.93 (0.66, 3.21) | 0.003 |
| Log-sTfR, mg/L* | SMA and NTS | 16 | 46.2 (36.5, 58.6) | Reference | | Reference | |
| | SMA | 33 | 42.8 (35.7, 51.4) | -0.08 (-0.39, 0.24) | 0.63 | -0.09 (-0.42, 0.24) | 0.59 |

| | | | | | | | |
|---|---|---|---|---|---|---|---|
| | NTS | 32 | 40.6 (32.6, 50.4) | -0.13 (-0.45, 0.18) | 0.41 | -0.15 (-0.48, 0.19) | 0.38 |
| | CM | 33 | 33.9 (29.8, 40.0) | -0.31 (-0.62, 0.004) | 0.05 | -0.30 (-0.62, 0.03) | 0.08 |
| Log-CRP, mg/L | SMA and NTS | 16 | 98.4 (72.9, 132.9) | Reference | | Reference | |
| | SMA | 33 | 96.7 (76.6, 122.0) | -0.02 (-0.57, 0.54) | 0.95 | -0.27 (-0.89, 0.36) | 0.40 |
| | NTS | 30 | 77.2 (51.7, 115.1) | -0.24 (-0.81, 0.32) | 0.39 | -0.32 (-0.95, 0.32) | 0.32 |
| | CM | 33 | 76.2 (51.6, 112.7) | -0.26 (-0.81, 0.30) | 0.36 | -0.49 (-1.11, 0.13) | 0.11 |
| Log-parasite density, parasites/µl | SMA and NTS | 16 | $9.1 \times 10^3$ ($3.0 \times 10^3$, $2.8 \times 10^4$) | Reference | | Reference | |
| | SMA | 33 | $5.1 \times 10^4$ ($2.4 \times 10^4$, $1.1 \times 10^4$) | 1.71 (0.56, 2.87) | 0.004 | 2.11 (0.84, 3.38) | 0.001 |
| | CM | 33 | $1.9 \times 10^5$ ($1.1 \times 10^5$, $3.2 \times 10^5$) | 3.01 (1.86, 4.17) | <0.001 | 3.31 (2.07, 4.57) | <0.001 |

Abbreviations: SMA, severe malaria anaemia; NTS, non-typhoidal *Salmonella* bacteremia; sTfR, soluble transferrin receptors; CRP, and C-reactive protein.

¶Adjusted coefficients (adj. β) and P-values were derived from a multivariable linear regression model adjusting for age in years, sex and underweight.

*Twelve sTfR values were above the upper limit of the assay (>84 mg/L) and were recorded as 84 mg/L for this analysis. These values were distributed across the groups as follows: SMA and NTS (2), SMA (5), NTS (2) and CM (3).

**Supplementary Table 4.** Correlation of log-hepcidin with iron and inflammatory biomarkers in hospitalized children

| | All groups | | SMA and NTS | | SMA | | NTS | | CM | |
|---|---|---|---|---|---|---|---|---|---|---|
| **Variable** | r | P | r | P | r | P | r | P | r | P |
| Log ferritin | 0.35 | 0.0002 | 0.14 | 0.60 | 0.10 | 0.60 | 0.66 | 0.0001 | 0.19 | 0.33 |
| Log sTfR | -0.36 | 0.0001 | -0.12 | 0.67 | -0.44 | 0.01 | -0.31 | 0.09 | -0.39 | 0.03 |
| Log CRP | 0.32 | 0.0005 | 0.04 | 0.89 | 0.55 | 0.0009 | 0.48 | 0.008 | 0.35 | 0.05 |
| Hemoglobin | 0.25 | 0.008 | 0.26 | 0.33 | 0.31 | 0.08 | -0.11 | 0.52 | 0.10 | 0.58 |
| Log parasitemia | 0.41 | 0.0001 | -0.09 | 0.73 | 0.51 | 0.002 | | | 0.27 | 0.12 |

Abbreviations: SMA, severe malaria anemia; NTS, non-typhoidal *Salmonella* bacteremia; CM, cerebral malaria; sTfR, soluble transferrin receptors; and CRP, C-reactive protein; and r, pairwise Pearson correlation coefficients.

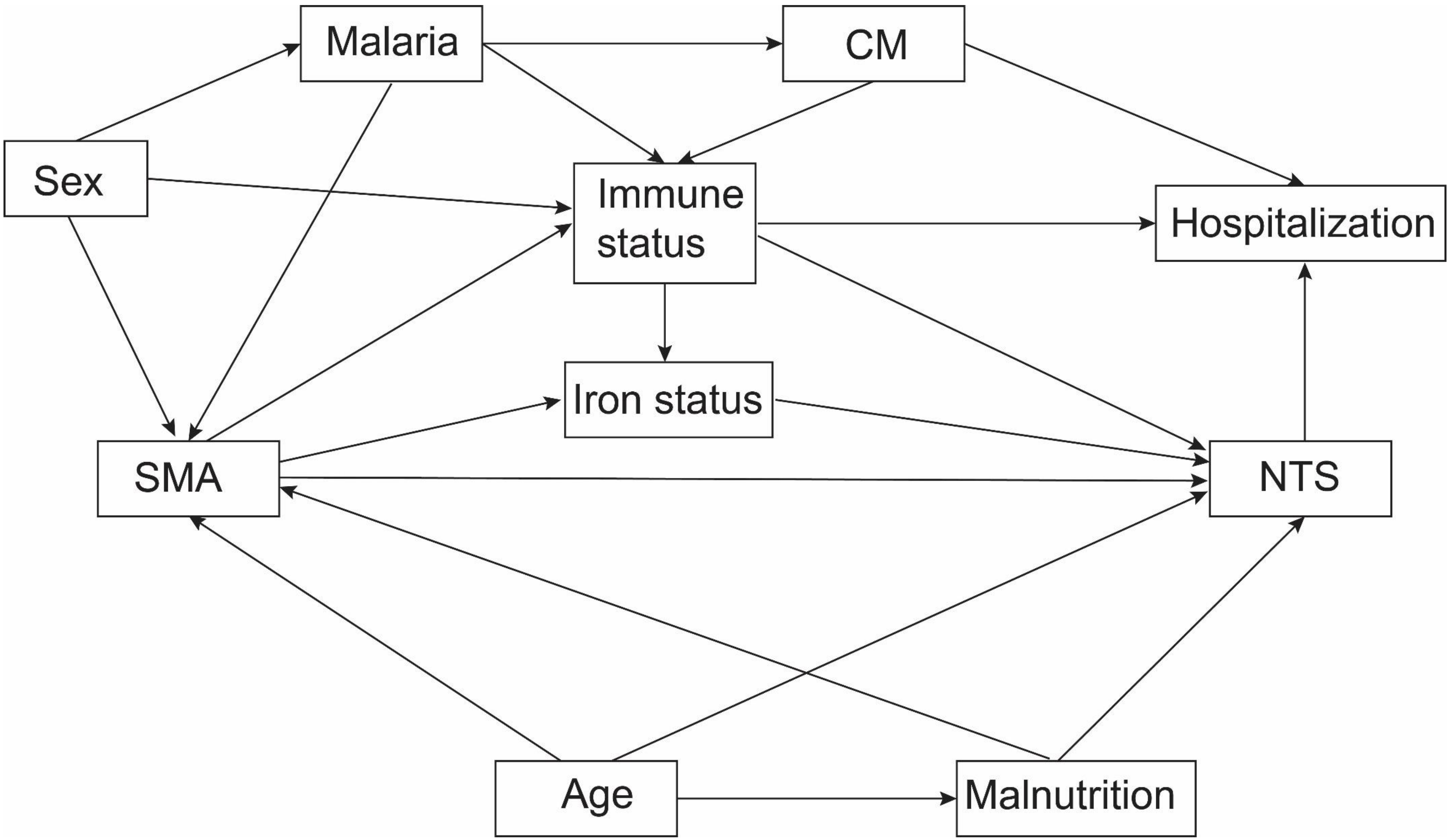


**Supplementary Figure 1**. Directed acyclic graph for the causal pathways between malaria and non-typhoidal *Salmonella* (NTS). CM denotes cerebral malaria; and SMA, severe malaria anemia.
